# Disentangling the relationship between fibromyalgia and insomnia: A bidirectional two-sample Mendelian random analysis

**DOI:** 10.1101/2024.10.28.24316268

**Authors:** Jin Gao, Liangke Pan, Zhenglun Pan

## Abstract

**Objective:** Fibromyalgia (FM) and insomnia frequently coexist, but whether FM is an independent causal risk factor for insomnia, and/or vice versa, remains unclear. We investigated the bidirectional causal relationship between FM and insomnia.

**Methods:** Genome-wide association studies (GWAS) summary statistics on FM [sample size = 416,757 (2689 cases; 299,606 controls) (FinnGen)] and insomnia [sample size = 462,341 (UK Biobank)] were used. The possible causal relationships were assessed by bi-directional MR analysis. The major analysis method of MR was performed using inverse-variance weighted (IVW), supplemented by MR-Egger, weighted median, weighted mode and simple mode approaches. In addition, sensitivity analyses including Cochran’s Q test, MR-Egger intercept test, and the leave-one-out test were conducted to ensure the stability of the results.

**Results:** There was strong evidence to suggest insomnia is a risk factor for FM (odds ratio (OR) 7.65, 95% confidence interval (CI) 3.17-18.45, p = 5.85E-06). We found no evidence that genetic liability to FM increases insomnia (OR = 1.00, 95% CI 0.99-1.01, p = 0.99)

**Conclusions:** This study provides evidence in support of a causal relationship between insomnia and increased risk of FM, whilst overcoming the major limitations of previous epidemiological studies. Interventions for insomnia may be an effective strategy to prevent or reduce the burden of FM. Extended study needs to be performed with multi-center, large-scale GWAS cohort studies to re-evaluate the association between FM and insomnia in future.

## Introduction

Fibromyalgia is a common disorder whose cardinal manifestation is chronic, widespread pain[1]. It affects approximately 2% to 4%of the general population [2]. More recent research characterizing it as a disorder of pain regulation and central sensitization[1]. Brain imaging studies using functional magnetic resonance imaging and other studies have shown that people with FM suffer from a number of disturbances in pain processing and regulation that amplify pain or reduce pain inhibition[3]. These changes may extend to processing of other sensory input, potentially explaining other bothersome symptoms, such as sleep disruption[4].

Insomnia is the term used to describe the unwelcome experience of having difficulty falling asleep. People describe insomnia as when they want and have the opportunity to sleep, but have trouble falling asleep or staying asleep or falling back asleep after waking up during the night, when they wake up earlier than expected, and/or when they feel that their sleep is not refreshing[5]. Insomnia is the most common sleep problem in population surveys [6]and is associated with chronic mental and physical health conditions[7-9]. In a longitudinal, community-based study of Norwegian women, insomnia symptoms approximately doubled the risk for new-onset FM[10]. It is suggested that there is a bidirectional relationship between FM and insomnia[5]. Although experts recommend that all patients with FM should receive sleep education to relieve pain, fatigue, and cognitive symptoms[1]. However, it is still unclear whether there is a causal relationship between FM and insomnia.

Mendelian randomization (MR) analysis is a research strategy for assessing causal relationships[11]. It uses genetic variants as instrumental variables (IVs), which obtained from genome-wide association studies (GWAS), to yield unconfounded information on the causal relationship between exposure and outcome and minimizes the limitations of observational studies. The cardinal principle of MR is genetic variants are randomly assigned during meiosis[12]; thus, they can be considered hereditary randomized controlled trials (RCTs) and may not be affected by residual confounding and reverse causality[13]. Here, we exploit large-scale GWAS data using two-sample MR to assess the putative bidirectional causality between FM and insomnia.

## Materials and methods

### Ethics

This study was reported according to STROBE-MR guidelines[14]. Deidentified summary data were collected from public GWAS which sought informed consent from their study participants. Ethical approval was not necessary.

### Study design

A bi-directional MR study was conducted to investigate the causal associations between FM and insomnia. Basically,MR analysis is subject to three assumptions: (I) the IVs are closely related to exposure (“relevance”); (II) the IVs are independent of any potential confounding factor (“exchangeability”); the IVs only affect outcome via the exposure (“exclusion restriction”). SNPs are used as IVs to determine the causal effect of exposure variables for MR analysis. The framework is described in Fig. 1.

**Fig. 1.**
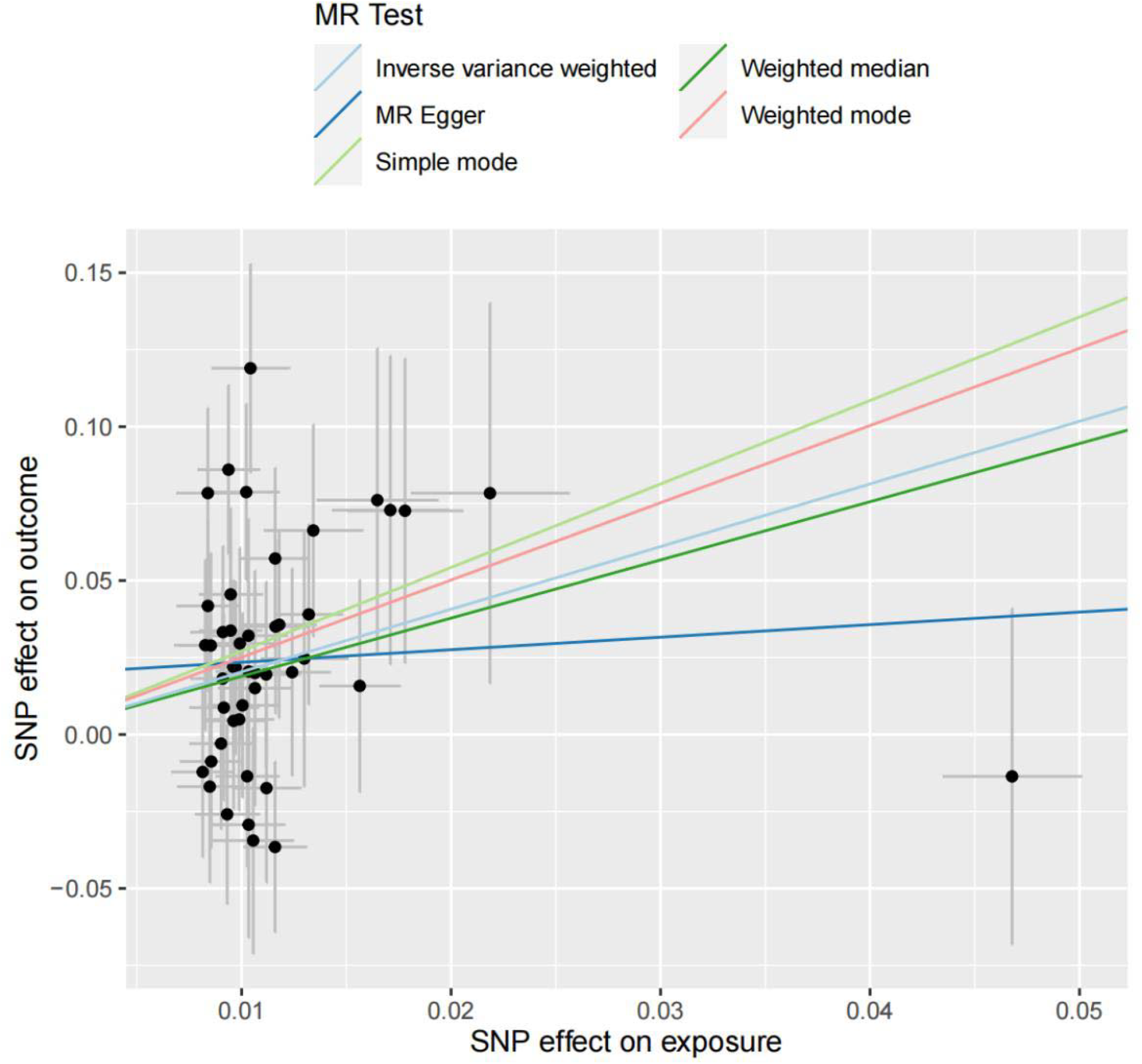
Scatter plot of the causal effect of insomnia on FM

### Data sources for FM and insomnia

The summary data of FM were obtained from the GWAS study from FinnGen(https://www.finngen.fi/en/) with the phenocode “finngen_R10_M13_FM” which consisted of 302,295 samples (2689 cases; 299,606 controls), and a total of 21,303,254 SNPs were genotyped at the FinnGen.

The summary data of insomnia were obtained from the GWAS study at the UK Biobank, including 462,341 individuals of European ancestry. a total of 9,851,867 SNPs were genotyped.

The information about each data source is provided in Table 1.

**Table 1.** Description of GWAS used for each phenotype.

| Phenotype | Data sources | Sample size | SNPs (n) | Ancestry |
| --- | --- | --- | --- | --- |
| FM | FinnGen | 302,295 | 21,303,254 | European |
| Insomnia | UK Biobank | 462341 | 9,851,867 | European |

### Selection of SNPs as IVs

SNPs with p-value<5e-8 were screened from GWAS data as preliminary IVs. After that, linkage disequilibrium (LD) analysis was performed on selected SNPs to ensure their independence (LD-r2<0.001 and clumping distance>10,000 kb). Since the main assumption of the MR analysis is that IVs can only affect the outcome through exposure, we eliminated SNPs associated with confounders using PhenoScanner (http://phenoscanner.medschl.cam.ac.uk/).

### Testing IV strength and statistical power

The strength of the IV was assessed using the F-statistic to minimize any possible weak IV bias. A higher F-statistic indicates a smaller bias[15].The following formula F = R^2^×(n−2)/1−R^2^(n: sample size of the GWAS; R^2^ : the proportion of explained variance of the IV) was used to calculate F-value[16]. And if F>10, it indicates that the study had sufficient strength. R^2^ was calculated by the formula: R^2^ = 2 × β^2^ × (1 − EAF) × EAF (β: estimate of the genetic effect of each SNP on iron status; EAF: effect allele frequency). The statistical power of MR analysis is calculated based on the variance explained by genetic instruments for the exposure,the proportion of cases, and the sample size of GWAS.

### Bidirectional MR analysis

Inverse variance weighted method (IVW), weighted median method, MR-Egger regression method, simple mode method and weighted mode method were used for MR Analysis of two samples to infer causality. IVW was used as the main analysis method for MR. IVW combined the MR Effect estimation of each SNP to obtain the overall weighted estimate of potential causal effect. When horizontal pleiotropy does not exist in instrumental variables, IVW analysis results are the most reliable [17]. Even if up to 50% of the information comes from the genetic variation of the invalid instrumental variable, the weighted median method can still obtain a consistent estimate of the causal effect[18]. The MR-Egger regression method can confirm whether there is horizontal pleiotropy in instrumental variables, and the estimated effect value of horizontal pleiotropy is represented by intercept. When horizontal pleiotropy exists in instrumental variables, the MR-Egger regression method can still obtain an unbiased estimate of causality[19]. If more than 50% of the information is derived from valid IVs, the weighted median method can give a valid causal estimate[19]. However, the power of the weighted median and MR-Egger methods which tend to provide wider confidence intervals (CI) are limited when compared to IVW [17], and are employed in this study only as complementary methods. Weighted mode and simple mode are two methods under the mode-based estimate approach, which is robust to horizontal pleiotropy. While the weighted mode exhibit higher precision compared to the simple mode, it shows an increased susceptibility to bias resulting from inside assumption violations[20].

### Sensitivity analyses

IV heterogeneity was examined by using the fixed effects model(P>0.05) or random effects model(P<0.05) according to Cochran’s Q statistic [34]. Horizontal pleiotropy was determined by conducting the MR-Egger intercept test. The outlier test was used to compares expected and observed distributions of each variant to identify outlier variants. The leave-one-out method was performed in the sensitivity analysis. The measure of the effects was odds ratios (ORs) and its 95% CI.

MR analyses were conducted using R version 4.3.1 with the “TwoSampleMR” (version 0.5.9) R packages. It was considered significant if the two-sided P-value was less than 0.05.

## Results

### Characteristics of the selected SNPs

SNPs strongly correlated with insomnia were extracted as IVs in GWAS(P<5e−8). After that, the LD analysis was performed (LD-r 2<0.001, clumping distance>10,000 kb) and palindromic variants resulting in potential strand ambiguity were removed. In addition, the PhenoScanner database was searched for risk factors. Eventually, 49 SNPs were enrolled in the MR analysis of insomnia on FM.

For the IVs of FM, only a small number (n=3) of SNPs were obtained when a strict P-value (P<5e−8) was taken for screening. To include more SNPs associated with FM, a more lenient threshold was adopted in this study (P<5e−6). After combining LD analysis and searching the PhenoScanner database for risk factors, 27 SNPs were obtained for the MR analysis of FM on insomnia finally.

The F-statistics for IVs were all greater than 10, thus IVs were generally considered to provide sufficient information for MR studies (Supplementary Tables 1 and 2).

### Causal effects of insomnia on FM risks

The results of this MR analysis are shown in Table 2 and Fig. 2. The OR of insomnia associated with FM for each of the 5 methods (IVW, weighted median, Weighted mode, MR-Egger and Simple mode) were 7.65(95% CI:3.17 to 18.45, P=5.85E-06),6.62(95% CI:2.00 to 21.88, P=0.002),12.30(95% CI:1.32 to 114.17, P=0.03),1.50(95% CI:0.10 to 21.88, P=0.77), and 15.07 (95% CI:0.98 to 232.15,P=0.06).Our results of the 5 methods showed that genetically predicted insomnia was significantly associated to the risk of FM (all P<0.05).

**Table 2.** Causal effects of insomnia on FM risks in MR analysis.

| Exposure | Outcome | SNPs (n) | MR method | OR | 95% CI | P-value |
| --- | --- | --- | --- | --- | --- | --- |
| Insomnia | FM | 49 | IVW | 7.65 | ( 3.17 , 18.45 ) | 5.85E-06 |
|  |  |  | Weighted median | 6.62 | ( 2.00 , 21.88 ) | 0.002 |
|  |  |  | Weighted mode | 12.30 | ( 1.32 , 114.17 ) | 0.03 |
|  |  |  | MR Egger | 1.50 | ( 0.10 , 21.88 ) | 0.77 |
|  |  |  | Simple mode | 15.07 | ( 0.98 , 232.15 ) | 0.06 |

**Fig. 2.**
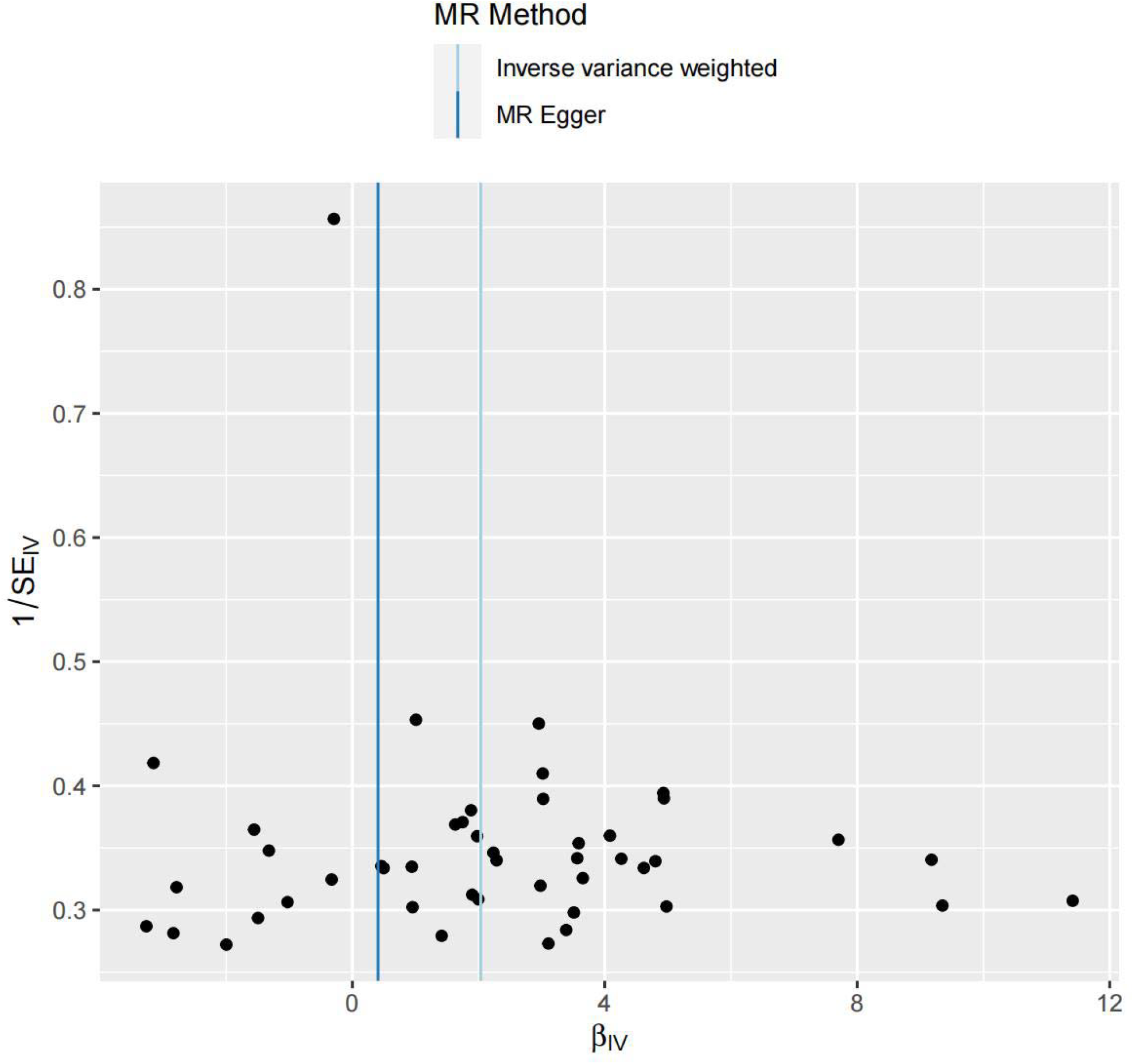
Scatter plot of the causal effect of FM on insomnia

### Causal effects of FM on insomnia

In Table 3 and Fig. 3, The OR of FM associated with insomnia for each of the 5 methods (IVW, weighted median, Weighted mode, MR-Egger and Simple mode) were 1.00(95% CI:0.99 to 1.01, P=0.99),1.00(95% CI:0.99 to 1.01, P=0.99),1.00(95% CI:0.99 to 1.01, P=0.93),1.00(95% CI:0.99 to 1.01, P=0.91),and 1.00(95% CI:0.99 to 1.01, P=0.76).Our results of the 5 methods showed that genetically predicted FM were not significantly associated to the risk of insomnia (all P>0.05).

**Table 3.**
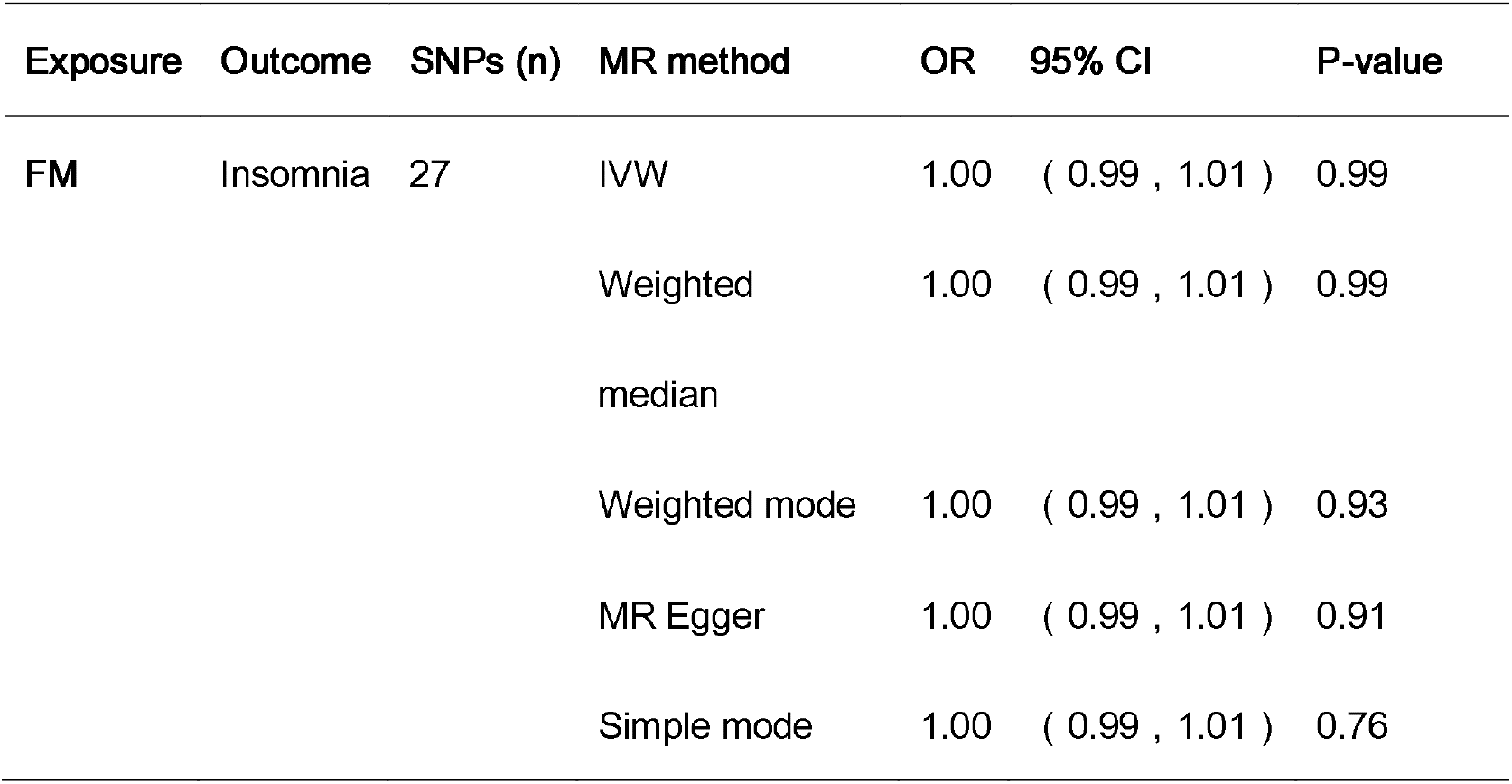
Causal effects of FM on insomnia risks in MR analysis.

**Table 4.**
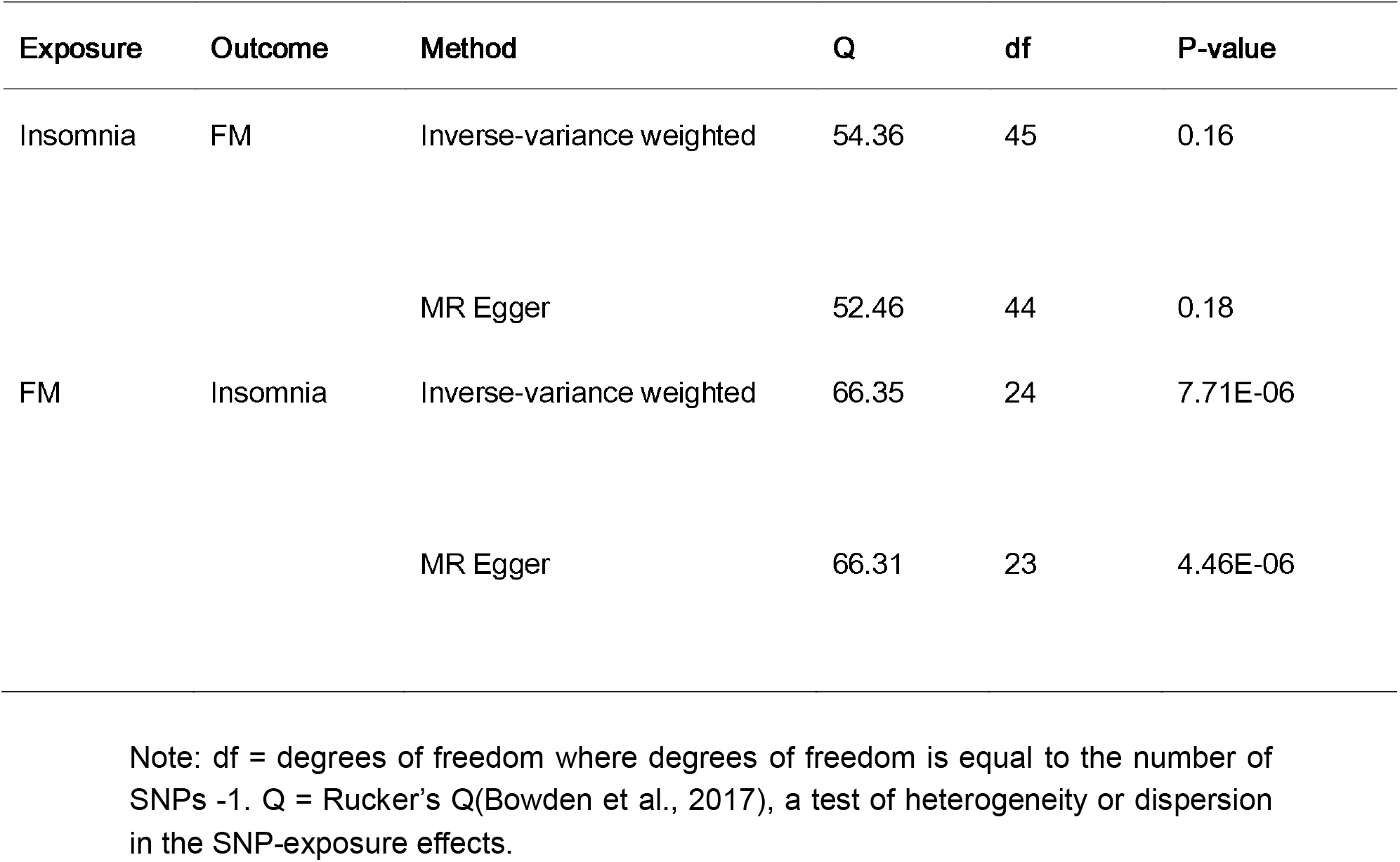
Tests of Heterogeneity in the SNP-exposure association.

**Table 5.**
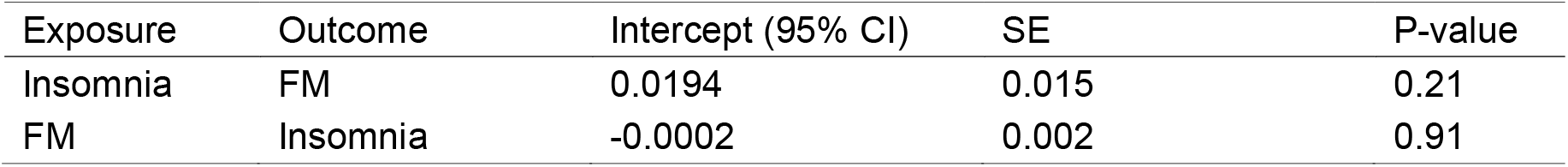
MR Egger test of directional pleiotropy.

| Exposure | Outcome | Intercept (95% CI) | SE | P-value |
| --- | --- | --- | --- | --- |
| Insomnia | FM | 0.0194 | 0.015 | 0.21 |
| FM | Insomnia | -0.0002 | 0.002 | 0.91 |

**Fig.3.**
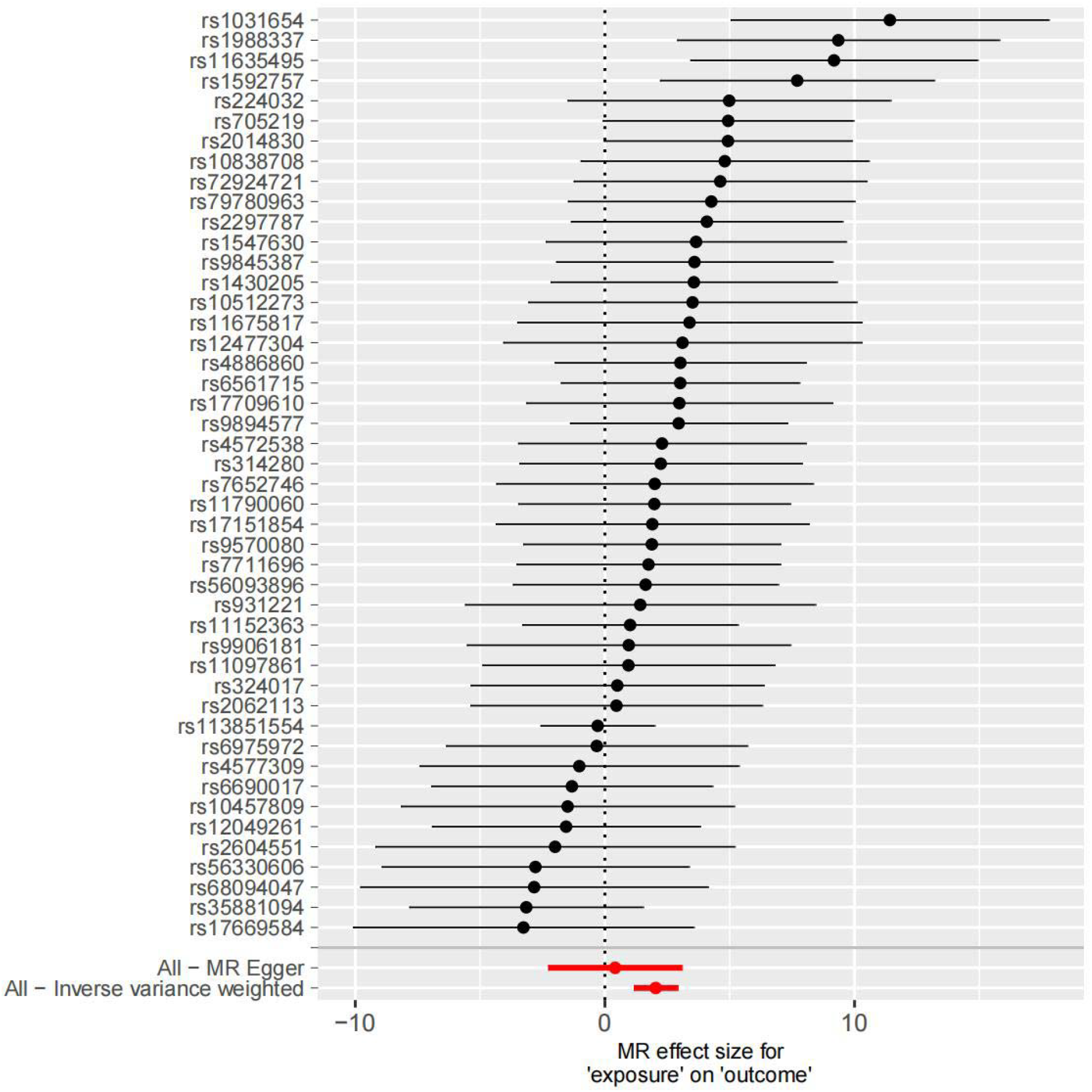
Leave-one-out sensitivity analysis of the effect of insomnia on FM

### Sensitivity analyses of MR

As MR Egger’s Q test was not statistically significant (Q=52.46, P=0.16), heterogeneity analysis on causal effects of insomnia on FM showed that there was no heterogeneity between the data. However, MR Egger’s Q test was statistically significant (Q=66.31, P=4.46E-06) in the opposite direction.

No directional pleiotropy bias was evident. The leave-one-out analysis demonstrated there was no SNP outliers, which suggests that our results were stable (see Fig. 4).

**Fig.4.**
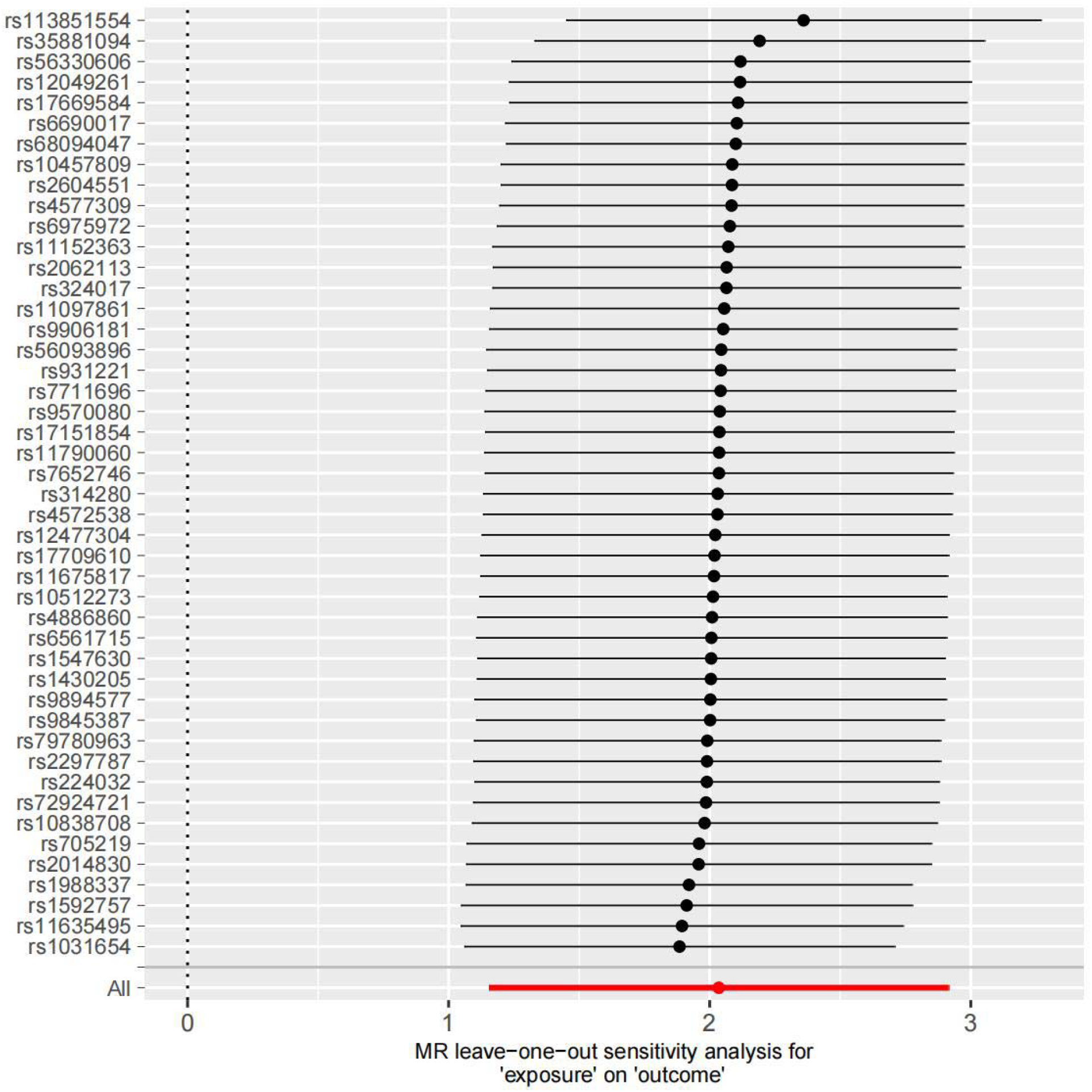
Leave-one-out sensitivity analysis of the effect of FM on insomnia

## Discussion

To our knowledge, this is the first bidirectional MR design to assess the causal effects between insomnia and FM. We found insomnia has causal effects on the pathogenesis of FM, however, forward MR revealed no evidence of causal effect of FM on insomnia.

The result of reverse MR was supported by many of the existing observational studies. Insomnia can be detected in 75% of the patients with FM[21]. A dose dependent association between poor sleep and FM symptom severity has been found in an epidemiological study[22]. Patients with FM with poor sleep quality were reported to show a higher degree of pain, poorer psychological status, more impaired body functions, and poorer quality of life than those with good sleep quality[23]. The effect of sleep on FM was further supported by results of clinical trials of pharmacotherapy which revealed Improved sleep quality can reduce pain in patients with FM[24, 25]. Pathophysiological research showed complex and multidimensional mechanisms underlie the effects of poor sleep on FM symptom severity (ie, myalgia, tenderness, and fatigue)[26, 27]. The current analysis suggested the causal risks to on FM from insomnia was genetically predicted.

The forward MR revealed no evidence of causal effect of FM on insomnia which contradicts some of the existing observational studies. For example, compared with healthy controls, most patients with FM report their sleep as poor or nonrestorative and feel unrefreshed upon awakening[23, 28]. Objective sleep quality as measured by polysomnography also supported that FM patients experienced shorter sleep duration, more time in light sleep, lower sleep efficiency, and longer duration of wakefulness during sleep than healthy individuals[29]. Our MR results might be explained by the fact that insomnia disorder is common in people with comorbid mental or physical health conditions affecting sleep, such as anxiety, depression, neurological and gastrointestinal disorder, however, the proportion of patients with FM is relatively small, the genetic predictive power from FM is offset by other diseases. In addition, although the F-statistics for IVs were all over 10, suggesting IVs were generally considered to provide sufficient information for MR studies, only a small number (n=3) of SNPs were obtained when a strict P-value (Pe<5−8) was taken for screening, and a more lenient threshold was used in this study (P<5e −6). Thus, the statistical power of IVs extracted from the FM GWAS dataset was insufficient to detect causation.

The strength of MR methods is to minimize residual confounding and reverse causality in observational studies. In the current study: MR assumptions were strictly followed: All the F-statistic of IVs were greater than 10; SNPs associated with outcomes were removed; SNPs associated with potential confounders were excluded. Finally, multiple sensitivity tests demonstrated the reliability of the findings.

There are some limitations that cannot be addressed at present. First, while FM and insomnia have their own diagnostic criteria[30, 31], they are also accompanied by many other clinical conditions, such as inflammatory diseases, psychological disorders, etc. If the study population includes a large number of people with secondary diagnoses, especially those with the same comorbidities or primary diseases, it is more difficult to distinguish confounders. Of note, we could not make it clear how many FM patients were included in the insomnia dataset and vice versa. Second, no weak instrumental variable bias exists in this study because the F-statistics were all > 10; however, only a small number (n=3) of SNPs were obtained when a strict P-value (P<5e−8) was taken for screening in reverse MR as IVs which generated an impetus to perform studies with larger sample size. Third, we should be cautious about generalizing the findings to other populations as the GWAs dataset is for populations of European descent. Extended study needs to be performed in future with improved GWAS database provided with detailed individualized sample information, etc.

## Conclusions

We found genetic evidence in support of a causal association between insomnia and FM risk. Interventions for insomnia may be an effective strategy to prevent or reduce the burden of FM. The underlying relationship between FM and insomnia is complex and worthy of further investigation. Extended study needs to be performed with multi-center, large-scale GWAS cohort studies to re-evaluate the association between FM and insomnia in future. Contributions The original idea for the study came at a family dinner when the daughter(Liangke Pan) asked her mother(Jin Gao), a psychologist, about the consequences of insomnia. The mother replied that insomnia has many serious consequences, and then said that healthy sleep is especially important for teenagers. Dad(Zhenglun Pan) said that fibromyalgia in rheumatology is related to sleep disorders, and improving sleep can ease the symptoms of fibromyalgia. The daughter further asked if there was a causal relationship between the two clinical questions, and the father replied that it was difficult to establish a causal relationship between diseases in medicine because it was against ethics to conduct disease-inducing tests on humans. Through a web search, Liangke Pan found that the two-sample Mendelian randomization method could answer causality between diseases, but required genome-wide association study data. So the family began the study.

All authors(family members) made contributions to the conception and design of the study and to the interpretation of the data. JG,LKP,ZLP were involved in the design of the study, the collection and analysis of data, and the drafting of the paper. JG and ZLP are responsible for data collection and analysis. ZLP also contributed to the manuscript. All authors read and approved the final manuscript.

## Data Availability

All data produced in the present study are available upon reasonable request to the authors

## Patient consent for publication

No data that could identify single patients are presented therefore this consent is not needed.

## Ethics approval and consent to participate

This study was reported according to STROBE-MR guidelines. Deidentified summary data were collected from public GWAS which sought informed consent from their study participants. Ethical approval was not necessary.

