## Supplementary Material for "Disentangling the relationship between fibromyalgia and insomnia: A bidirectional two-sample Mendelian random analysis"

**Supplementary Table 1. Summary of genetic variants (n=49) used to estimate the effect of insomnia on FM in MR analyses.**

| SNP | Chr | Pos (GRCh37) | Effect allele | Other allele | EAF | Beta | Se | P | R^2^ | F |
| --- | --- | --- | --- | --- | --- | --- | --- | --- | --- | --- |
| rs1031654 | 13 | 53807900 | A | C | 0.798844 | -0.0104245 | 0.00185446 | 1.90E-08 | 6.83E-05 | 31.59902251 |
| rs10457809 | 6 | 100536524 | T | C | 0.475096 | 0.00814109 | 0.0014875 | 4.40E-08 | 6.48E-05 | 29.95361782 |
| rs10512273 | 9 | 100842269 | G | A | 0.505024 | -0.00826645 | 0.00148454 | 2.60E-08 | 6.71E-05 | 31.0064753 |
| rs10838708 | 11 | 47419962 | A | G | 0.45899 | -0.00947656 | 0.00150271 | 2.90E-10 | 8.60E-05 | 39.76941491 |
| rs11097861 | 4 | 104408976 | G | A | 0.716256 | 0.0100437 | 0.00164946 | 1.10E-09 | 8.02E-05 | 37.07677992 |
| rs11152363 | 18 | 55389957 | A | G | 0.186319 | 0.0156393 | 0.00192518 | 4.50E-16 | 0.000142714 | 65.99181298 |
| rs113851554 | 2 | 66523432 | T | G | 0.057291 | 0.0467802 | 0.00331329 | 2.90E-45 | 0.000430979 | 199.3440926 |
| rs11635495 | 15 | 67512344 | C | T | 0.512179 | 0.00937342 | 0.00148509 | 2.80E-10 | 8.62E-05 | 39.83719286 |
| rs11675817 | 2 | 58941908 | A | G | 0.381698 | 0.00853076 | 0.00152888 | 2.40E-08 | 6.73E-05 | 31.13341568 |
| rs11790060 | 9 | 93440650 | C | T | 0.330834 | -0.0103391 | 0.00157854 | 5.80E-11 | 9.28E-05 | 42.89951777 |
| rs12049261 | 1 | 106647785 | C | G | 0.292534 | 0.0111868 | 0.00163048 | 6.80E-12 | 0.000101806 | 47.07376304 |
| rs12477304 | 2 | 200105648 | G | T | 0.202918 | -0.0103283 | 0.00184766 | 2.30E-08 | 6.76E-05 | 31.24724236 |
| rs1430205 | 5 | 88382768 | T | C | 0.461507 | 0.00947493 | 0.00149107 | 2.10E-10 | 8.73E-05 | 40.37886216 |
| rs1547630 | 13 | 111988214 | A | G | 0.651513 | 0.00910814 | 0.00156447 | 5.80E-09 | 7.33E-05 | 33.89401654 |
| rs1592757 | 5 | 104554297 | C | G | 0.355788 | 0.0102216 | 0.00154996 | 4.30E-11 | 9.41E-05 | 43.49055041 |
| rs17151854 | 8 | 10379049 | T | G | 0.15241 | 0.0129893 | 0.00207434 | 3.80E-10 | 8.48E-05 | 39.21116375 |
| rs17669584 | 17 | 30572596 | G | A | 0.195283 | 0.0105512 | 0.00191225 | 3.40E-08 | 6.58E-05 | 30.44475564 |
| rs17709610 | 10 | 102490521 | G | A | 0.297979 | -0.0099161 | 0.00162078 | 9.50E-10 | 8.10E-05 | 37.4310301 |
| rs1988337 | 4 | 90371049 | G | A | 0.552395 | 0.00838726 | 0.00149645 | 2.10E-08 | 6.79E-05 | 31.41332535 |
| rs2014830 | 3 | 50134964 | T | C | 0.303519 | -0.0116018 | 0.0016233 | 8.90E-13 | 0.00011047 | 51.08004757 |
| rs2062113 | 16 | 59442275 | C | T | 0.568257 | -0.00961678 | 0.00150275 | 1.60E-10 | 8.86E-05 | 40.95283861 |
| rs224032 | 10 | 62762069 | A | G | 0.550358 | 0.00839066 | 0.00149108 | 1.80E-08 | 6.85E-05 | 31.66565515 |
| rs2297787 | 10 | 102920380 | A | T | 0.080193 | -0.0178001 | 0.00274996 | 9.60E-11 | 9.06E-05 | 41.89770669 |
| rs2604551 | 4 | 15089577 | G | T | 0.640384 | -0.00848084 | 0.00155238 | 4.70E-08 | 6.45E-05 | 29.84556352 |
| rs2644128 | 1 | 201824312 | G | C | 0.548279 | 0.0106284 | 0.00149103 | 1.00E-12 | 0.000109889 | 50.81139746 |
| rs2803296 | 1 | 1916670 | C | G | 0.543586 | -0.00862206 | 0.00149047 | 7.30E-09 | 7.24E-05 | 33.46368201 |
| rs314280 | 6 | 104952962 | G | A | 0.547047 | 0.00971363 | 0.00149134 | 7.30E-11 | 9.18E-05 | 42.42363739 |
| rs324017 | 12 | 57094031 | C | A | 0.705433 | -0.00988248 | 0.00163145 | 1.40E-09 | 7.94E-05 | 36.69293681 |
| rs35881094 | 2 | 58695786 | G | T | 0.426882 | 0.0115952 | 0.00150555 | 1.30E-14 | 0.000128277 | 59.31496014 |
| rs4572538 | 2 | 146665444 | T | C | 0.364055 | -0.00960596 | 0.00156161 | 7.70E-10 | 8.18E-05 | 37.8385518 |
| rs4577309 | 2 | 190424107 | G | A | 0.533655 | -0.00854833 | 0.00149214 | 1.00E-08 | 7.10E-05 | 32.82022353 |
| rs4886860 | 15 | 74047995 | C | G | 0.767408 | -0.011796 | 0.00175566 | 1.80E-11 | 9.76E-05 | 45.14262612 |
| rs56093896 | 2 | 113346389 | A | C | 0.214053 | -0.0124111 | 0.00181352 | 7.70E-12 | 0.000101291 | 46.83537179 |
| rs56330606 | 19 | 37183051 | G | A | 0.378954 | 0.00930932 | 0.00153044 | 1.20E-09 | 8.00E-05 | 36.99997439 |
| rs6561715 | 13 | 53314391 | A | T | 0.630662 | -0.011623 | 0.00154234 | 4.80E-14 | 0.000122818 | 56.79032485 |
| rs6690017 | 1 | 57377057 | G | T | 0.408855 | -0.0102669 | 0.00151022 | 1.10E-11 | 1.00E-04 | 46.21642524 |
| rs68094047 | 12 | 109417396 | T | C | 0.251267 | 0.0103356 | 0.0017167 | 1.70E-09 | 7.84E-05 | 36.24771993 |
| rs6975972 | 7 | 1030832 | G | A | 0.578726 | -0.00902066 | 0.00150428 | 2.00E-09 | 7.78E-05 | 35.95980984 |
| rs705219 | 3 | 117983423 | A | T | 0.887373 | 0.013423 | 0.00235299 | 1.20E-08 | 7.04E-05 | 32.54296647 |
| rs72924721 | 11 | 65818519 | T | C | 0.073137 | 0.016478 | 0.00288103 | 1.10E-08 | 7.07E-05 | 32.71233591 |
| rs7652746 | 3 | 49224874 | G | A | 0.62362 | -0.00909657 | 0.00153206 | 2.90E-09 | 7.62E-05 | 35.25347278 |
| rs7711696 | 5 | 136150847 | T | G | 0.305042 | 0.0111716 | 0.00161138 | 4.10E-12 | 0.000103951 | 48.06544174 |
| rs79780963 | 10 | 103192742 | T | C | 0.077385 | -0.0170989 | 0.00277578 | 7.30E-10 | 8.21E-05 | 37.94585839 |
| rs8180817 | 7 | 114407487 | C | G | 0.431016 | -0.0100553 | 0.00150974 | 2.70E-11 | 9.59E-05 | 44.35921607 |
| rs931221 | 12 | 84327328 | A | T | 0.236738 | 0.0106361 | 0.00175313 | 1.30E-09 | 7.96E-05 | 36.8073628 |
| rs9570080 | 13 | 59258641 | C | T | 0.344131 | -0.0106379 | 0.00157858 | 1.60E-11 | 9.82E-05 | 45.41263856 |
| rs9845387 | 3 | 116707088 | A | C | 0.040279 | -0.0218562 | 0.00377648 | 7.10E-09 | 7.24E-05 | 33.49446579 |
| rs9894577 | 17 | 45145925 | A | G | 0.3182 | 0.0132051 | 0.00159687 | 1.30E-16 | 0.000147883 | 68.3820925 |
| rs9906181 | 17 | 21394374 | G | A | 0.687567 | -0.00915026 | 0.00163869 | 2.40E-08 | 6.74E-05 | 31.1796606 |

**Supplementary Table 2. Summary of genetic variants (n = 4) used to estimate the effect of FM on insomnia in MR analyses.**

| SNP | Chr | Pos (GRCh37) | Effect allele | Other allele | EAF | Beta | Se | P | R^2^ | F |
| --- | --- | --- | --- | --- | --- | --- | --- | --- | --- | --- |
| rs1004867 | 16 | 49601221 | A | G | 0.283978 | -0.138616 | 0.0302639 | 4.64E-06 | 6.94E-05 | 20.97848226 |
| rs11214585 | 11 | 113357001 | C | G | 0.441284 | -0.126554 | 0.0277113 | 4.95E-06 | 6.90E-05 | 20.85619504 |
| rs114507934 | 1 | 154851599 | C | T | 0.0632504 | 0.27714 | 0.0589583 | 2.59E-06 | 7.31E-05 | 22.09559224 |
| rs11588064 | 1 | 224051206 | T | C | 0.207505 | 0.157993 | 0.0343812 | 4.32E-06 | 6.99E-05 | 21.11692884 |
| rs12342816 | 9 | 104616638 | C | T | 0.00686894 | -0.670762 | 0.146154 | 4.44E-06 | 6.97E-05 | 21.06263089 |
| rs12429980 | 13 | 45976003 | A | C | 0.391269 | 0.132898 | 0.0283587 | 2.78E-06 | 7.26E-05 | 21.96146869 |
| rs12457428 | 18 | 45152381 | T | C | 0.323382 | -0.140488 | 0.0290473 | 1.32E-06 | 7.74E-05 | 23.39182177 |
| rs1338221 | 1 | 219627563 | C | T | 0.825015 | 0.165596 | 0.0356922 | 3.49E-06 | 7.12E-05 | 21.52534769 |
| rs147764067 | 16 | 17963371 | G | T | 0.0197427 | -0.451887 | 0.0924184 | 1.01E-06 | 7.91E-05 | 23.90781744 |
| rs17050273 | 2 | 59583489 | A | G | 0.171339 | -0.16933 | 0.035948 | 2.47E-06 | 7.34E-05 | 22.18786309 |
| rs17559134 | 1 | 223858602 | A | G | 0.308098 | -0.137625 | 0.0294928 | 3.07E-06 | 7.20E-05 | 21.775081 |
| rs2327966 | 20 | 15802583 | C | T | 0.268545 | -0.162548 | 0.0306233 | 1.11E-07 | 9.32E-05 | 28.17451244 |
| rs34323745 | 7 | 23272449 | A | C | 0.245331 | 0.157893 | 0.0325783 | 1.26E-06 | 7.77E-05 | 23.48908196 |
| rs347908 | 1 | 119640976 | T | A | 0.429351 | 0.129601 | 0.0279209 | 3.46E-06 | 7.13E-05 | 21.54542203 |
| rs4661271 | 1 | 223492066 | A | G | 0.452282 | -0.144915 | 0.0275371 | 1.42E-07 | 9.16E-05 | 27.69410954 |
| rs4715124 | 6 | 13143886 | A | G | 0.574685 | -0.134232 | 0.0280029 | 1.64E-06 | 7.60E-05 | 22.97752407 |
| rs56132815 | 2 | 104258841 | C | T | 0.157243 | -0.187929 | 0.0369482 | 3.65E-07 | 8.56E-05 | 25.87010317 |
| rs56307343 | 8 | 64187989 | C | T | 0.0231451 | -0.393505 | 0.0852808 | 3.95E-06 | 7.04E-05 | 21.2909532 |
| rs6045664 | 20 | 1959499 | C | T | 0.933009 | -0.282222 | 0.057478 | 9.10E-07 | 7.97E-05 | 24.10879816 |
| rs61951132 | 13 | 20332185 | C | T | 0.00653643 | -0.703093 | 0.14969 | 2.64E-06 | 7.30E-05 | 22.06160467 |
| rs62375296 | 5 | 120075684 | C | T | 0.0259758 | -0.435741 | 0.0806181 | 6.48E-08 | 9.66E-05 | 29.21385445 |
| rs7078231 | 10 | 82653650 | A | T | 0.58688 | -0.139076 | 0.0280572 | 7.16E-07 | 8.13E-05 | 24.57043546 |
| rs7327894 | 13 | 61087963 | C | G | 0.180036 | 0.169923 | 0.036583 | 3.40E-06 | 7.14E-05 | 21.57460368 |
| rs76155771 | 12 | 115358281 | G | A | 0.0187766 | 0.507154 | 0.109821 | 3.87E-06 | 7.05E-05 | 21.32583522 |
| rs77266689 | 3 | 122903543 | T | C | 0.0282859 | -0.357471 | 0.0780491 | 4.65E-06 | 6.94E-05 | 20.97697971 |
| rs78243804 | 6 | 111239862 | C | T | 0.0694697 | 0.26245 | 0.0566799 | 3.65E-06 | 7.09E-05 | 21.44036281 |
| rs7910831 | 10 | 15507081 | T | G | 0.201048 | -0.160658 | 0.0337783 | 1.97E-06 | 7.48E-05 | 22.62175331 |

Note: SNP, single nucleotide polymorphism; Chr, chromosome; EAF, effect allele frequency; SE, standard error.
